# Genetic associations of eosinophilic granulomatosis with polyangiitis in the Japanese population: Exploring similarities and differences with European populations

**DOI:** 10.64898/2025.12.10.25341817

**Authors:** Aya Kawasaki, Ikue Ito-Naito, Mina Sato, Yuka Kawamura, Ken-ei Sada, Kenji Itoh, Nobuyuki Ono, Fumio Hirano, Tomoaki Higuchi, Shigeto Kobayashi, Kenji Nagasaka, Takahiko Sugihara, Takashi Fujimoto, Makio Kusaoi, Yuya Kondo, Kumiko Shimoyama, Keita Yamashita, Takashi Higuchi, Hajime Kono, Noriyoshi Ogawa, Isao Matsumoto, Kunihiro Yamagata, Takayuki Sumida, Hiroshi Hashimoto, Hirofumi Makino, Yoshihiro Arimura, Masayoshi Harigai, Naoto Tamura, Naoyuki Tsuchiya

## Abstract

**Objectives:** Eosinophilic granulomatosis with polyangiitis (EGPA) is classified as a subtype of anti-neutrophil cytoplasmic antibody (ANCA)-associated vasculitis (AAV). A recent genome-wide association study (GWAS) of EGPA in populations of European ancestry reported several susceptibility loci. In view of population differences in genetic factors of AAV, we made an attempt to examine the contribution of these loci to EGPA in a Japanese population.

**Methods:** The EGPA associated variants in *LPP*, *TSLP*, *IRF1*/*IL5*, *HLA*, *CDK6*, 10p14 and 12q21 in European populations were analyzed in a total of 103 Japanese patients with EGPA, including 48 ANCA-positive and 45 ANCA-negative patients. Allele frequency data on approximately 60,000 Japanese individuals from the Japanese Multi Omics Reference Panel (jMorp) served as the controls.

**Results:** An increasing tendency in *TSLP* rs1837253 C allele was observed in total EGPA, myeloperoxidase (MPO)-ANCA-positive EGPA and ANCA-negative EGPA. In addition, association of *HLA-DRB1*07:01* was observed in total EGPA and MPO-ANCA-positive EGPA. These findings supported the previous findings in the European populations. On the other hand, unlike in the European populations, association of *DRB1*09:01* was observed in Japanese EGPA. With respect to amino acid residues in HLA-DRβ1, Val 78 encoded by *DRB1*07:01* and \**09:01* was associated with total EGPA and MPO-ANCA-positive EGPA with genome-wide significance (P<5.0E-08). Association with the other loci was not observed in Japanese EGPA.

**Conclusion:** *TSLP* was suggested to be associated with EGPA both in the European and Japanese populations regardless of the ANCA status. In *HLA-DRB1*, both shared and population-specific susceptibility alleles were observed.

**What is already known on this topic:**

- Susceptibility genes to eosinophilic granulomatosis with polyangiitis (EGPA) have been reported by a genome-wide association study (GWAS) in populations of European ancestry.
- In non-European populations, GWAS of EGPA has not been reported and genetic factors contributing to the development of EGPA are largely unknown.

**What this study adds:**

- **TSLP* rs1837253C and HLA-DRB1*07:01 were shown to be associated with EGPA both in European and Japanese populations.*
- *HLA-DRB1*09:01* was shown to be associated with EGPA in the Japanese, but not in the European populations.

**How this study might affect research, practice or policy:**

- This study showed that *TSLP* is a shared susceptibility gene to EGPA in the European and Japanese populations, implicating its significance as a molecular target as well as a potential biomarker for response to IL-5 targeted therapy across populations.
- Population differences are observed in allelic associations other than *TSLP*, which should be taken into consideration in the process of drug and biomarker development for EGPA.

## INTRODUCTION

Eosinophilic granulomatosis with polyangiitis (EGPA) is a subset of anti-neutrophil cytoplasmic antibody (ANCA)-associated vasculitis (AAV), characterized by asthma and eosinophilia.^1, 2^ The prevalence of ANCA positivity ranges from 30% to 47%.^2^ It is reported that clinical features differ between ANCA-positive and -negative EGPA patients.^1, 2^ The incidence of EGPA is low both in Europe and Japan, with annual incidence of 0.9/million in UK, and 2.4/million in Japan.^3^ Partly due to its low incidence, genetic factors of EGPA remain largely unclear.

Genome-wide association studies (GWAS) on AAV have been reported in European populations.^4–7^ One of them focused on EGPA and reported 11 susceptibility loci, many of which were also associated with asthma or increased eosinophil count.^7^ Among the associated loci, *BCL2L11*, *TSLP* and 10p14 showed an association with EGPA regardless of the ANCA status. On the other hand, *HLA-DQ* was specifically associated with myeloperoxidase (MPO)-ANCA-positive EGPA, and *IRF1*/*IL5* and *GPA33* were associated with ANCA-negative EGPA. These results suggested genetic differences between ANCA-positive and -negative EGPA.

GWAS on AAV in the Asian populations has not been published thus far. However, candidate gene studies suggest substantial difference between genetic factors for AAV between European and East Asian populations.^8^ We previously reported an association of *HLA-DRB1*09:01*-*DQA1*03:02*-*DQB1*03:03* haplotype with susceptibility to MPO-ANCA-positive AAV, ^9,10^ which includes about a half of Japanese patients with EGPA. However, statistically significant association of *HLA-DRB1* alleles with EGPA *per se* was not detected,^9^ probably due to low statistical power.

In this study, we focused on the EGPA associated genes reported in the GWAS on European populations,^7^ and examined whether they are also associated with EGPA in the Japanese population.

## METHODS

### Patients and controls

Japanese patients with AAV were recruited at institutes affiliated with Japan Research Committee of the Ministry of Health, Labour, and Welfare for Intractable Vasculitis (JPVAS), Research Committee of Intractable Renal Disease organized by the Ministry of Health, Labour and Welfare, Japan, as well as research groups organized by University of Tsukuba, Tokyo Medical and Dental University and Tokyo Women’s Medical University. All patients were Japanese and living in Japan. Genomic DNA from the patients was collected between 1999 and 2024. All available samples at the time of analysis were included in the study. The AAV patients were classified into 427 patients with microscopic polyangiitis (MPA), 161 with granulomatosis with polyangiitis (GPA), 103 with EGPA, and 40 with unclassifiable AAV, based on the European Medicines Agency (EMA) algorism (Table 1).^11^ Among the EGPA patients, 48 were MPO-ANCA-positive and 45 were ANCA-negative. The other 10 patients include those who were MPO-ANCA-negative/PR3-ANCA-positive, and those with missing data on ANCA status.

**Table 1.** Characteristics of the patients with ANCA-associated vasculitis.

| Category | Number |
| --- | --- |
| ANCA-associated vasculitis (AAV) | 731 (450) |
| Sex: Male / Female | 285 / 446 |
| Classification according to the EMA algorithm |  |
| Microscopic polyangiitis (MPA) | 427 (273) |
| Granulomatosis with polyangiitis (GPA) | 161 (88) |
| Eosinophilic granulomatosis with polyangiitis (EGPA) | 103 (57) |
| Unclassifiable | 40 (32) |
| Classification according to ANCA specificity |  |
| MPO-ANCA-positive AAV | 587 (363) |
| MPO-ANA-positive non-EGPA | 539 (334) |
| PR3-ANCA-positive AAV | 92 (55) |
| EGPA | 103 (57) |
| Sex: Male / Female | 36 / 67 |
| Subtype |  |
| MPO-ANCA-positive EGPA | 48 (29) |
| ANCA-negative EGPA | 45 (25) |
| *others | 10 (3) |

\*Others include patients who are MPO-ANCA negative/PR3-ANCA positive, and those with missing data on ANCA status.
The numbers in parentheses indicate the number of overlapping samples with the previous study.<sup>9</sup>
EMA: European Medicines Agency

The patients with EGPA were analyzed for association with the EGPA associated variants in the European GWAS^7^ in a case-control study. With respect to *TSLP* rs1837253 and *HLA-DRB1* alleles, patients with MPA, GPA and unclassifiable AAV were also genotyped, to address the specificity of association with EGPA. Data from 60KJPN consisting of approximately 60,000 Japanese individuals, available from the Japanese Multi Omics Reference Panel (jMorp), were used as the control data.^12, 13^

### Genotyping

The single nucleotide variants (SNVs) rs9290877 in *LPP*, rs1837253 in *TSLP*, rs11745587 in *IRF1*/*IL5*, rs42041 in *CDK6* and rs78478398 in 12q21 were genotyped using the rhAmp SNP Genotyping System (Integrated DNA Technologies, Inc. Coralville, IA, USA). The primers used in this assay are listed in Supplementary Table S1. As for rs9274704 (A/G) in *HLA-DQ* and rs34574566 (T-repeat) in 10p14, genotyping was performed using Sanger sequencing, because of incompatibility of the rhAmp SNP Genotyping System with a repeat variant (rs34574566), or due to the presence of nearby variants (rs9274704). Cycle sequencing was performed using the SupreDye v3.1 kit (Edge Biosystems, Inc. San Jose, CA, USA), and the products were subsequently sequenced on an Applied Biosystems 3500 Genetic Analyzer (Thermo Fisher Scientific, Waltham, MA, USA). Primers used in the sequencing analysis and representative sequencing chromatogram in each genotype of rs9274704 and rs34574566 are shown in Supplementary Table S1 and Supplementary Figure S1, respectively. To overcome the difficulty in accurately genotyping the number of repeat nucleotides of homopolymer by Sanger sequencing, a mismatched base (T to A) was introduced into the forward primer for rs34574566.

Genotyping of *HLA-DRB1* allele was performed using WAKFlow HLA typing kit (Wakunaga Pharmaceutical Co., Ltd., Osaka, Japan) in 528 patients and long-read sequencing in 37 patients. In the long-read sequencing analysis, *HLA-DRB1* gene was amplified using the NGSgo-AmpX v2 HLA-DRB1 Whole Gene kit (GenDx, Utrecht, The Netherlands) and the amplicons were sequenced using the Sequel IIe system (PacBio, Menlo Park, CA, USA). *HLA-DRB1* alleles were determined by the NGSengine version 2.31.0 (GenDx) software using fastq data obtained from long-read sequencing. One of the EGPA patients analyzed using long-read sequencing was excluded from statistical analysis, because *HLA-DRB1* allele could not be determined; thus the number of the EGPA patients analyzed for *HLA* was102. For each amino acid position of HLA-DRβ1, allele count was determined based on HLA amino acid sequences deposited in the IPD-IMGT/HLA Database.^14^

### Statistical analysis

Association was tested using chi-squared test or Fisher’s exact test using a 2×2 contingency table under the allelic model. Fisher’s exact test was used when any cell in the table contained a count of five or less. Allele frequencies were compared between each AAV subset (total EGPA, MPO-ANCA-positive EGPA, ANCA-negative EGPA, MPA, GPA, MPO-ANCA-positive AAV, MPO-ANCA-positive non-EGPA and PR3-ANCA-positive AAV) and 60KJPN. Correction for multiple testing was performed using the false discovery rate (FDR) method.^15^ The significance level was set at 0.05 for corrected P value (P_c_). The number of multiple tests used for correction is shown in each table.

### Ethics statement

This study was approved by the ethics committees of University of Tsukuba, Tokyo Women’s Medical University, Tokyo Medical and Dental University and all other participating institutions. This study was performed in accordance with the Declaration of Helsinki. Written Informed consent was obtained from all participants.

## RESULTS

### Association studies on the European EGPA-associated variants in a Japanese population

Association of the European EGPA-associated variants^7^ was analyzed in Japanese patients with EGPA. Data from 60KJPN in jMorp database served as control data.^12, 13^ Associations of these variants were examined in total EGPA, MPO-ANCA-positive EGPA and ANCA-negative EGPA. The results are shown in Table 2.

**Table 2.** Association analysis of the European EGPA associated variants in the Japanese patients with EGPA.

| Variant ID-Effect allele | Gene/<br>region | Total EGPA (n=103) |  |  |  | MPO-ANCA-positive EGPA (n=48) |  |  |  | ANCA-negative EGPA (n=45) |  |  |  | 60KJPN<br>(controls) |
| --- | --- | --- | --- | --- | --- | --- | --- | --- | --- | --- | --- | --- | --- | --- |
|  |  | n (EAF) | P<sub>uncorr</sub> | P<sub>c</sub> | OR (95%CI) | n (EAF) | P<sub>uncorr</sub> | P<sub>c</sub> | OR (95%CI) | n (EAF) | P<sub>uncorr</sub> | P<sub>c</sub> | OR (95%CI) | n (EAF) |
| rs9290877-C | *LPP* | 7 (0.034) | 0.232 | 0.57 | 1.58 (0.74-3.36) | 3 (0.031) | 0.469 | 0.644 | 1.45 (0.46-4.57) | 3 (0.033) | 0.450 | 0.644 | 1.55 (0.49-4.89) | 2612 (0.022) |
| rs1837253-C | *TSLP* | 90 (0.437) | 0.00484 | 0.058 | 1.48 (1.13-1.95) | 43 (0.448) | 0.0314 | 0.218 | 1.55 (1.04-2.32) | 44 (0.489) | 0.00372 | 0.058 | 1.83 (1.21-2.76) | 41188 (0.344) |
| rs11745587-A | *IRF1/IL5* | 78 (0.379) | 0.445 | 0.644 | 1.12 (0.84-1.48) | 33 (0.344) | 0.847 | 0.920 | 0.96 (0.63-1.46) | 35 (0.389) | 0.479 | 0.644 | 1.17 (0.76-1.78) | 42341 (0.353) |
| rs9274704-G | *HLA-DQ* | 97 (0.471) | 0.189 | 0.57 | 1.20 (0.91-1.58) | 51 (0.531) | 0.0363 | 0.218 | 1.53 (1.02-2.28) | 39 (0.433) | 0.882 | 0.920 | 1.03 (0.68-1.57) | 51018 (0.426) |
| rs42041-G | *CDK6* | 4 (0.019) | 0.823 | 0.92 | 0.77 (0.29-2.07) | 4 (0.042) | 0.308 | 0.592 | 1.69 (0.62-4.60) | 0 (0) | 0.178 | 0.570 | - | 3005 (0.025) |
| rs78478398-C | 12q21 | 13 (0.063) | 0.720 | 0.864 | 1.11 (0.63-1.94) | 7 (0.073) | 0.510 | 0.644 | 1.29 (0.60-2.79) | 3 (0.033) | 0.492 | 0.644 | 0.57 (0.18-1.79) | 6867 (0.057) |
| rs34574566 | 10p14 |  |  |  |  |  |  |  |  |  |  |  |  |  |
| (T)<sub>8</sub> |  |  |  |  |  |  |  |  |  |  |  |  |  | 8 (0.00007) |
| (T)<sub>12</sub> |  |  |  |  |  |  |  |  |  |  |  |  |  | 2 (0.00002) |
| (T)<sub>13</sub> |  |  |  |  |  |  |  |  |  |  |  |  |  | 351 (0.003) |
| (T)<sub>14</sub> |  | 128 (0.621) |  |  | referent | 59 (0.615) |  |  | referent | 57 (0.633) |  |  | referent | 80907 (0.675) |
| (T)<sub>15</sub> |  | 54 (0.262) | 0.152 | 0.570 | 1.26 (0.92-1.73) | 25 (0.260) | 0.321 | 0.592 | 1.27 (0.79-2.02) | 25 (0.278) | 0.257 | 0.570 | 1.31 (0.82-2.10) | 27059 (0.226) |
| (T)<sub>16</sub> |  | 24 (0.117) | 0.219 | 0.570 | 1.31 (0.85-2.03) | 12 (0.125) | 0.261 | 0.570 | 1.42 (0.77-2.65) | 8 (0.089) | 0.964 | 0.964 | 0.98 (0.47-2.06) | 11549 (0.096) |
| (T)<sub>17</sub> |  |  |  |  |  |  |  |  |  |  |  |  |  | 4 (0.00003) |

Data from 60KJPN, consisting of approximately 60,000 Japanese individuals and provided by the Japanese Multi Omics Reference Panel (jMorP), were used as control data.<sup>12, 13</sup>
n (EAF): effect allele number (effect allele frequency), OR: odds ratio, CI: confidence interval, P<sub>uncorr</sub>: uncorrected P, P<sub>c</sub>: P corrected using the false discovery rate method based on 24 statistical tests.<sup>15</sup>

*TSLP* rs1837253 C allele was increased in total EGPA, MPO-ANCA-positive EGPA and ANCA-negative EGPA, which was consistent with European GWAS.^7^ Although the association barely missed significance after correction for multiple comparisons, odds ratios (OR) in the Japanese population were comparable to those in the European GWAS (Supplementary Table S2). Similarly, a tendency toward association of *HLA-DQ* rs9274704 was observed also in Japanese MPO-ANCA-positive EGPA.

On the other hand, significant association with rs9290877 in *LPP*, rs11745587 in *IRF1*/*IL5*, rs42041 in *CDK6*, and rs78478398 in 12q21 was not observed in Japanese EGPA. The variant rs34574566, located in 10p14 region, consists of T repeats ranging from 8 to 26 repeats. In the Japanese population, (T)_14_ is the most frequent allele, followed by (T)_15_ and (T)_16_. When the frequencies of (T)_15_ and (T)_16_ were compared with (T)_14_ as a reference, significant difference was not detected between Japanese EGPA and controls.

The SNVs rs72689399 in *GPA33*, rs72946301 in *BCL2L11* and rs6454802 in *BACH2* were excluded from the analysis due to their low allele frequency (minor allele frequency < 0.01) in the Japanese population. With respect to rs6931740 in the *HLA* region, significant deviation from Hardy–Weinberg equilibrium was detected (P value for Hardy–Weinberg equilibrium test: 2.06E-07). According to the jMorp,^12, 13^ a copy number variation is observed in the region surrounding rs6931740 (jMorp CNV ID: hg38-chr6-32679300). Therefore, this variant was also excluded from the analysis.

### Association study on *TSLP* rs1837253 in AAV subsets

To examine which AAV subsets are associated with *TSLP* rs1837253 in the Japanese population, association was also tested with MPA, GPA, MPO-ANCA-positive AAV, MPO-ANCA-positive non-EGPA, and proteinase 3 (PR3)-ANCA-positive AAV. As shown in Table 3, rs1837253 C allele showed a trend of increase in MPO-ANCA-positive AAV (uncorrected P [P_uncorr_]=0.040, OR 1.13, 95%confidence interval [CI] 1.01-1.28). On the other hand, the trend was marginal in MPO-ANCA-positive non-EGPA (P_uncorr_=0.132, OR 1.10, 95%CI 0.97-1.25) as compared with MPO-ANCA-positive EGPA (OR 1.55, 95%CI 1.04-2.32) and ANCA-negative EGPA (OR1.83, 95%CI 1.21-2.76) (Table 2). Taken together, these results may suggest the specificity of *TSLP* association with susceptibility to EGPA.

**Table 3.**
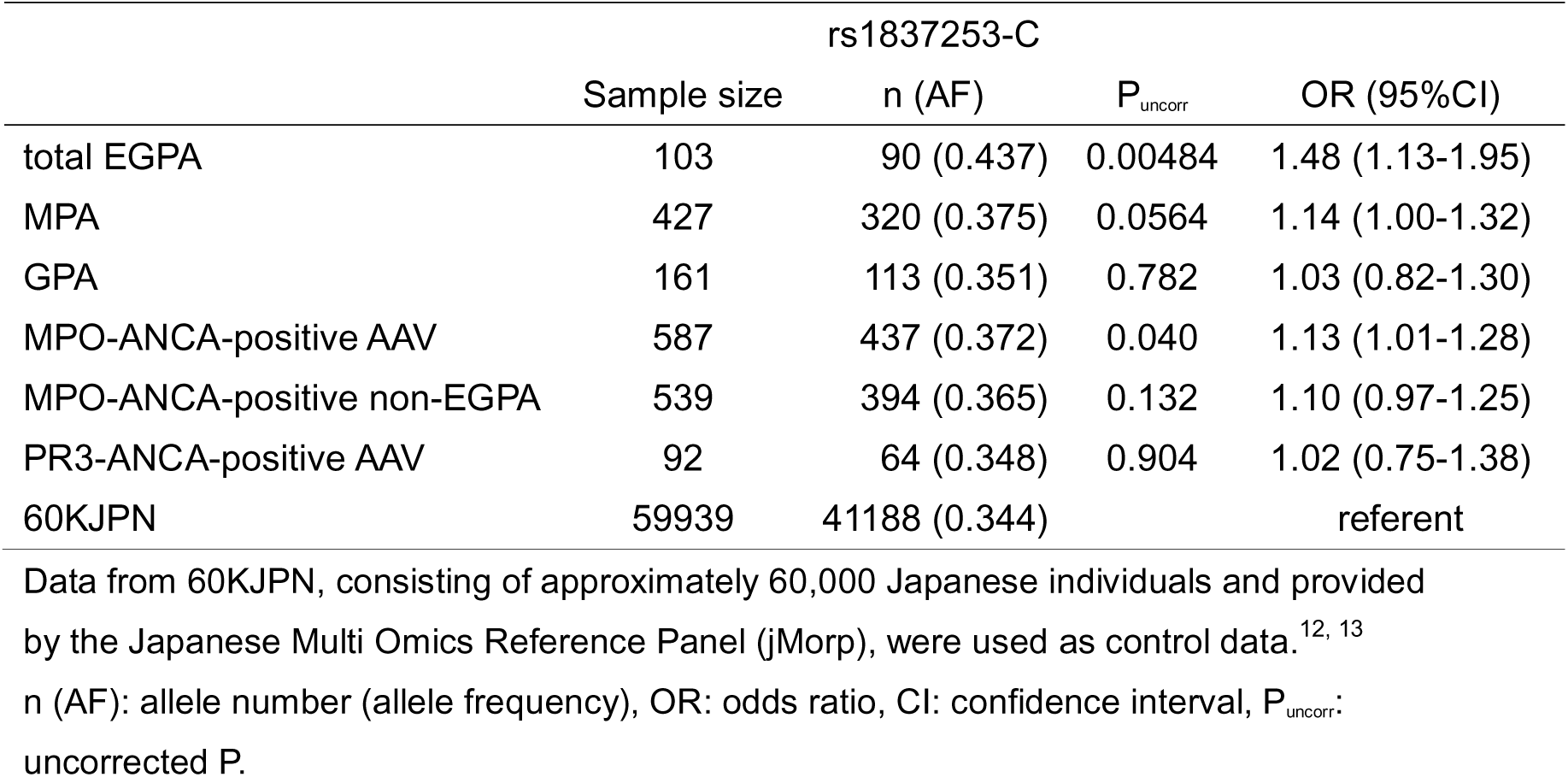
Association analysis on *TSLP* rs1837253 with AAV subsets.

### Association of *HLA* alleles and amino acids with EGPA

In our previous study with a smaller sample size of EGPA (56 patients), association of *HLA-DRB1* allele with EGPA did not reach statistical significance.^9^ However, in the present study with a larger sample size, a genome-wide significant association was observed between *HLA-DRB1*09:01* and MPO-ANCA-positive EGPA (Table 4). In addition, association of *DRB1*09:01* with total EGPA, and that of *DRB1*07:01* with total EGPA and MPO-ANCA-positive EGPA were detected (P_c_ < 0.05). When the association was tested in MPO-ANCA-positive non-EGPA patients, *DRB1*09:01* was significantly associated (P_uncorr_=5.84E-10, OR 1.70, 95%CI 1.44-2.02), whereas *DRB1*07:01* was not (P_uncorr_=0.501, OR 1.32, 95%CI 0.42-4.11) (Supplementary Table S3).

**Table 4.**
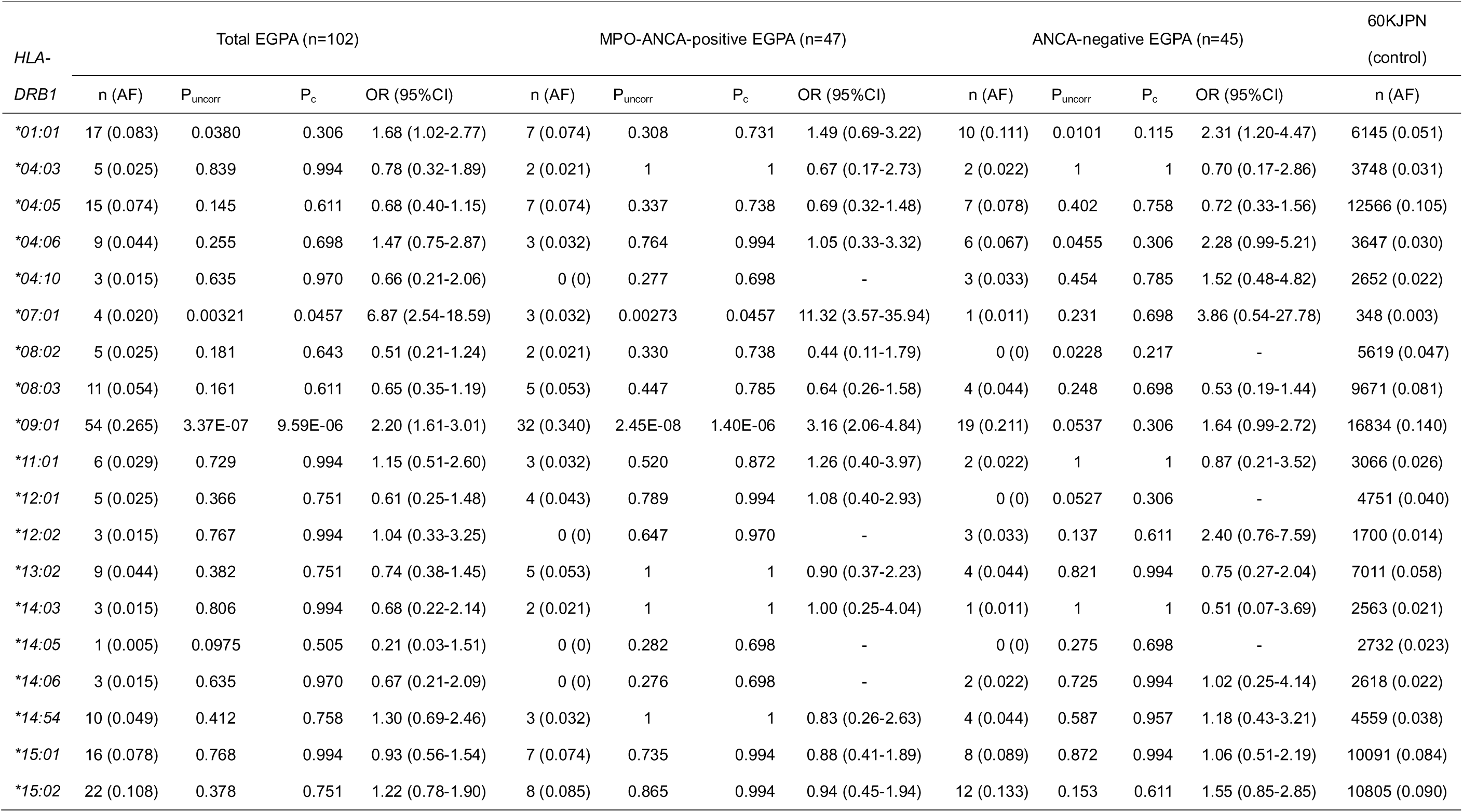
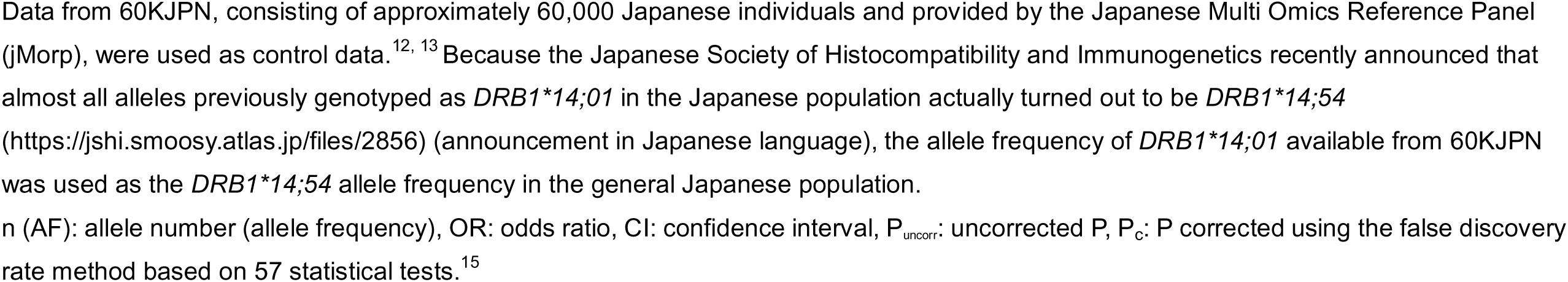
Association analysis of *HLA-DRB1* alleles with EGPA.

In the analysis of HLA-DRβ1 amino acids at each position, the strongest association was observed in Val at position 78 (Total EGPA: P_uncorr_=1.31E-08, OR 2.36, 95%CI 1.74-3.20, MPO-ANCA-positive EGPA: P_uncorr_=3.21E-10, OR 3.52, 95%CI 2.32-5.35), (Table 5 and Supplementary Table S4). Along with Val 78, eight amino acids of HLA-DRβ1 were associated with total EGPA and/or MPO-ANCA-positive EGPA with genome-wide significance. These amino acids are mainly encoded by *DRB1*07:01* and *\*09:01*; therefore, it was difficult to distinguish the associations of HLA amino acids from those of *HLA* alleles. As shown in Supplementary Table S4, associations of HLA-DRβ1 amino acids were also detected in ANCA-negative EGPA (P_c_ < 0.05).

**Table 5.** Genome-wide significant association of HLA-DRβ1 amino acids with EGPA.

| HLA-DRβ1 |  | Total EGPA (n=102) |  |  | MPO-ANCA-positive EGPA (n=47) |  |  | ANCA-negative EGPA (n=45) |  |  | 60KJPN<br>(control) | HLA-DRB1 allele<br>encoding the amino<br>acid |
| --- | --- | --- | --- | --- | --- | --- | --- | --- | --- | --- | --- | --- |
| position | amino<br>acid | n (AF) | P<sub>uncorr</sub> | OR (95%CI) | n (AF) | P<sub>uncorr</sub> | OR (95%CI) | n (AF) | P<sub>uncorr</sub> | OR (95%CI) | n (AF) |  |
| 4 | Q | 58 (0.284) | 4.27E-08 | 2.29 (1.69-3.10) | 35 (0.372) | 9.14E-10 | 3.42 (2.25-5.19) | 20 (0.222) | 0.0472 | 1.65 (1.00-2.71) | 17182 (0.148) | *07:01, *09:01 |
| 9 | K | 54 (0.265) | 4.96E-07 | 2.18 (1.60-2.98) | 32 (0.340) | 3.43E-08 | 3.13 (2.04-4.79) | 19 (0.211) | 0.0593 | 1.62 (0.98-2.69) | 16918 (0.142) | *09:01 |
| 11 | D | 54 (0.265) | 4.99E-07 | 2.18 (1.60-2.98) | 32 (0.340) | 3.44E-08 | 3.12 (2.04-4.79) | 19 (0.211) | 0.0594 | 1.62 (0.98-2.69) | 16920 (0.142) | *09:01 |
| 26 | Y | 55 (0.270) | 2.05E-07 | 2.22 (1.63-3.03) | 32 (0.340) | 3.98E-08 | 3.11 (2.03-4.77) | 20 (0.222) | 0.0302 | 1.72 (1.05-2.83) | 16989 (0.142) | *03:01, *09:01 |
| 28 | H | 54 (0.265) | 4.99E-07 | 2.18 (1.60-2.98) | 32 (0.340) | 3.44E-08 | 3.12 (2.04-4.79) | 19 (0.211) | 0.0594 | 1.62 (0.98-2.69) | 16920 (0.142) | *09:01 |
| 30 | G | 54 (0.265) | 4.99E-07 | 2.18 (1.60-2.98) | 32 (0.340) | 3.44E-08 | 3.12 (2.04-4.79) | 19 (0.211) | 0.0594 | 1.62 (0.98-2.69) | 16920 (0.142) | *09:01 |
| 31 | I | 71 (0.348) | 2.29E-08 | 2.23 (1.67-2.97) | 39 (0.415) | 5.45E-08 | 2.96 (1.96-4.46) | 29 (0.322) | 0.00197 | 1.98 (1.27-3.09) | 23073 (0.193) | *01:01, *09:01 |
| 78 | V | 58 (0.284) | 1.31E-08 | 2.36 (1.74-3.20) | 35 (0.372) | 3.21E-10 | 3.52 (2.32-5.35) | 20 (0.222) | 0.0354 | 1.69 (1.03-2.79) | 17217 (0.144) | *07:01, *09:01 |
| 181 | M | 58 (0.284) | 1.99E-07 | 2.20 (1.62-2.98) | 35 (0.372) | 3.62E-09 | 3.28 (2.16-4.99) | 20 (0.222) | 0.0684 | 1.58 (0.96-2.60) | 17783 (0.153) | *07:01, *09:01, *10:01 |

Data from 60KJPN, consisting of approximately 60,000 Japanese individuals and provided by the Japanese Multi Omics Reference Panel (jMorp), were used as control data.<sup>12, 13</sup>
n (AF): allele number (allele frequency), OR: odds ratio, CI: confidence interval, P<sub>uncorr</sub>: uncorrected P.

When linkage disequilibrium between the MPO-ANCA-positive EGPA associated variant, rs9274704, and amino acids of HLA-DRβ1 was examined, strong linkage disequilibrium was observed between rs9274704 and HLA-DRβ1 amino acids, Glu 98 and Ala 104 (*r^2^* = 0.833) (Supplementary Table S5). Glu 98 and Ala 104 are in absolute linkage disequilibrium (*r^2^* = 1) and are carried by *DRB1*04*, *\*07:01* and *\*09:01* alleles. Considering these findings, the slight association of rs9274704 in MPO-ANCA-positive EGPA may be attributable to that of *DRB1*07:01* and *\*09:01* alleles.

## DISCUSSION

The epidemiology of AAV is substantially different between the populations of European and Asian ancestries,^3^ which appears to be caused, at least in part, by the differences in the genetic background.^8^ Although a GWAS of EGPA has been reported on populations of European ancestry,^7^ it has not been reported on Asian populations. Furthermore, the European EGPA susceptibility variants have not been tested for replication in Asian populations. Thus, the genetic factors for EGPA remain largely unknown in Asian populations. In this study, we made an attempt to examine whether the European GWAS loci are also associated with EGPA in a Japanese population.

With respect to *HLA* loci, *DRB1*0801*-*DQA1*04:01*-*DQB1*04:02* and *DRB1*07:01*-*DQA1*02:01*-*DQB1*02:02*/*DQB1*03:03* haplotypes as well as *DRB1*01:03* allele have been reported to be associated with MPO-ANCA-positive EGPA in the European populations. In this study, we detected the association of *DRB1*07:01* with MPO-ANCA-positive EGPA, suggesting that the association of the *DRB1*07:01* haplotype may be shared between European and Japanese populations.

On the other hand, the frequencies of *DRB1*08:01* and *DRB1*01:03* are both extremely low in the Japanese population;^12,13^ in fact, these alleles were not detected in the EGPA patients in this study. In contrast, *DRB1*09:01*, previously reported to be associated with MPO-ANCA-positive AAV,^8–10^ was also shown to be associated with MPO-ANCA-positive EGPA in this study. Interestingly, *DRB1*09:01* is the most frequent *HLA-DRB1* allele in the general Japanese population, but rare in the European populations. Thus, these alleles may represent population-specific susceptibility alleles to EGPA.

In the major histocompatibility complex (*MHC*) region, European GWAS has reported the association of rs9274704, located at the intergenic region close to *HLA-DQB1,* with MPO-ANCA-positive EGPA.^7^ The present study also detected a similar trend of association in the Japanese population. The risk allele rs9274704 G was found to be in strong linkage disequilibrium with HLA-DRβ1 Glu 98 and Ala 104 *(r^2^*= 0.833) (Supplementary Table S5). Both amino acids are encoded by *DRB1*04*, *\*07:01* and *\*09:01*. Among these alleles, *DRB1*07:01* and *\*09:01* were increased in MPO-ANCA-positive EGPA (*\*07:01*: P_uncorr_=0.00273, OR 11.32, 95%CI 3.57-35.94, *\*09:01*: P_uncorr_=2.45E-08, OR 3.16, 95%CI 2.06-4.84) (Table 4). Thus, the increase of rs9274704 G is likely to be secondarily caused by the linkage disequilibrium with *DRB1*09:01* and *\*07:01*, at least in the Japanese population. We also elaborated on this issue in the European populations by leveraging the 1000 genomes project genotype data (European, EUR) of rs9274704 and *HLA-DRB1* alleles available from Ensembl (release 115) and the International Genome Sample Resource (IGSR).^16.17^ Linkage disequilibrium with rs9274704 was observed between both *DRB1*07:01 (D’*=0.918, *r^2^*=0.317) and *DRB1*08:01* (*D’*=0.889, *r^2^*=0.095). Therefore, it appears possible that the association of rs9274704 might be caused by linkage disequilibrium with *HLA* alleles; however, this issue needs to be confirmed using individual patient data.

When the association of each HLA-DRβ1 amino acid was analyzed, the strongest association was observed at Val 78, which constitutes the peptide-binding pocket 4 of HLA-DRβ1, and is thought to influence peptide binding affinity and specificity. As shown in Table 4, the amino acid residues in HLA-DRβ1 associated with MPO-ANCA-positive EGPA are mainly encoded by *DRB1*09:01* and *\*07:01*; therefore, it is difficult to determine whether specific amino acids, or the *HLA-DRB1* alleles as a whole, are primarily associated with MPO-ANCA-positive EGPA. The association with HLA-DRβ1 amino acids was also detected in ANCA-negative EGPA, although the association did not reach genome-wide significance. It should be noted that *HLA-DQ* variant was reported to be associated with ANCA-negative EGPA in the European populations;^7^ thus, HLA-class II appears to be associated with susceptibility not only to MPO-ANCA-positive EGPA, but also to ANCA-negative EGPA.

We previously reported the association of *DRB1*09:01* haplotype with MPO-ANCA-positive AAV including EGPA.^8–10^ As shown in Supplementary Table S3, the association of *DRB1*09:01* was also observed in MPO-ANCA-positive non-EGPA patients, but *DRB1*07:01* (P_uncorr_=0.501, OR 1.32, 95%CI 0.42-4.11) was not significantly associated. These observations may suggest that *DRB1*09:01* is associated with MPO-ANCA-positive AAV in general, not specifically with EGPA. On the other hand, *DRB1*07:01* may be specifically associated with MPO-ANCA-positive EGPA. Further studies with larger sample size may be required to test this hypothesis.

*TSLP* encodes thymic stromal lymphopoietin, a cytokine involved in type 2 inflammation and mainly secreted by epithelial cells.^18, 19^ TSLP induces Th2 cytokine production, including IL-4, IL-5 and IL-13, through multiple pathways. Dendritic cells, T helper 2 (Th2) cells, group 2 innate lymphoid cells (ILC2s), and mast cells have been reported to be involved in the Th2 cytokine production.^18,19^ IL-5 is essential for the differentiation, maturation, proliferation and activation of eosinophils and is implicated in the pathogenesis of EGPA.^19–21^ Monoclonal antibodies to IL-5 (mepolizumab) and IL-5 receptor α (benralizumab) are used for treatment of EGPA.^21, 22^ The SNV rs1837253 is located in an active enhancer region upstream of *TSLP* gene, suggesting a potential regulatory role. There is no variant in linkage disequilibrium (*r^2^* > 0.3) with rs1837253 in European populations (EUR) according to the 1000 genomes data (*r^2^* data were obtained from the LDlink).^23^ Therefore, it is likely that rs1837253 itself may be causally associated with pathogenesis. It was reported that TSLP secretion was higher in primary nasal epithelial cells (NECs) from individuals with rs1837253 C/C genotype compared to those with C/T or T/T genotype, under stimulation with poly I:C.^24^ This is in line with the findings that serum TSLP levels were upregulated in EGPA patients compared with healthy controls.^25^ However, significant association between rs1837253 and expression of *TSLP* gene was not reported in eQTL databases including the Genotype-Tissue Expression (GTEx) Portal, ImmuNexUT or eQTLGen Phase I.^26–28^ Thus, the mechanism by which rs1837253 influences TSLP secretion requires further study.

*TSLP* rs1837253 was also reported to be associated with asthma and increased eosinophil count.^7^ This variant may be associated with EGPA and asthma by inducing type 2 inflammation, which plays a role in the pathogenesis of the diseases.^19^ Lyons et al. suggested that eosinophilia caused by susceptibility genes including *TSLP* may contribute to the etiology of EGPA.^7^

This study has several limitations. First, the sample size was relatively small due to the rarity of EGPA, especially when patients were stratified according to ANCA status. One possible reason for the failure to replicate previously reported associations in European populations is the limited statistical power of this study due to the small sample size. Power to detect association of the European GWAS variants is shown in Supplementary Table S6.^29^ Most of the power values were below 0.8.

In this study, only the variants reported in the European GWAS were examined, except for *HLA-DRB1* gene. The possibility that other variants in the same set of genes might be associated in the Japanese population cannot be excluded. In order to overcome this issue, GWAS on Japanese AAV is underway.

It should be noted that in this study the AAV patients were classified according to the EMA algorism,^11^ rather than the ACR/EULAR 2022 criteria.^30–32^ The recruitment of the patients started in 1999, and the clinical data was insufficient for classification using the ACR/EULAR 2022 criteria. Sada et al. reported that 97.6% of the Japanese EGPA patients and 2.5% of AAV patients other than EGPA classified by the EMA algorism were reclassified into EGPA by the ACR/EULAR 2022 criteria.^33^ Thus, the association study results might be somewhat different when patients were reclassified according to ACR/EULAR 2022 criteria.

Genotype data in AAV patients were obtained using the rhAmp SNP Genotyping System, Sanger sequencing and WAKFlow HLA typing kit, while those in controls (60KJPN) were obtained using short-read sequencing. A possibility that differences in the genotyping methods may have affected the results cannot be fully excluded. With respect to *HLA-DRB1*09:01*, significant association with MPO-ANCA-positive EGPA was also detected when compared with the control data in our previously study obtained using WAKFlow HLA typing kit (P_uncorr_=2.54E-06, OR 2.86, 95%CI 1.82-4.51).^9^

In conclusion, our findings suggested that *TSLP* may contribute to disease susceptibility to EGPA in European and Japanese populations regardless of ANCA status. *HLA-DRB1* is strongly associated with MPO-ANCA-positive EGPA in both populations, with shared as well as ancestry-specific susceptibility alleles.

## Acknowledgements

The authors are grateful to all the patients for participating in this study. Part of this work was presented at the 66th Annual Scientific Meeting of the Japan College of Rheumatology (Modern Rheumatology 2022;32[Suppl]:S117).

## Contributors

AK and NTs designed the study, interpreted the data and wrote the manuscript. AK, MS, and YK performed genotyping; AK also conducted statistical analysis, and II-N performed *HLA* sequencing. K-ES, KuY, HM, YA, MH and NTa coordinated the cohorts.

KeY coordinated the patient recruitment at Hamamatsu University Hospital. K-ES, KI, NOn, FH, ToH, SK, KN, TSug, TF, MK, YK, KS, TaH, HK, NOg, IM, TSum and HH recruited the participants and collected clinical data. All authors contributed to the article and approved the submitted version.

## Patient consent for publication

Not applicable.

## Ethics approval

This study was approved by the University of Tsukuba Institute of Medicine Ethics Committee (approval ID: 122(3), 268, 180, 227, G303), as well as by all the institutes participating in this study. This study was conducted in accordance with the Ethical Guidelines for Medical and Biological Research Involving Human Subjects. Written informed consent was obtained from all participants.

## Data availability statement

The data supporting the findings of this study are included in the article or in supplementary materials. The genotype data are not publicly available to protect participant privacy.

