## Supplementary Figure for "Genetic associations of eosinophilic granulomatosis with polyangiitis in the Japanese population: Exploring similarities and differences with European populations"

Content

Supplementary Figure S1

A

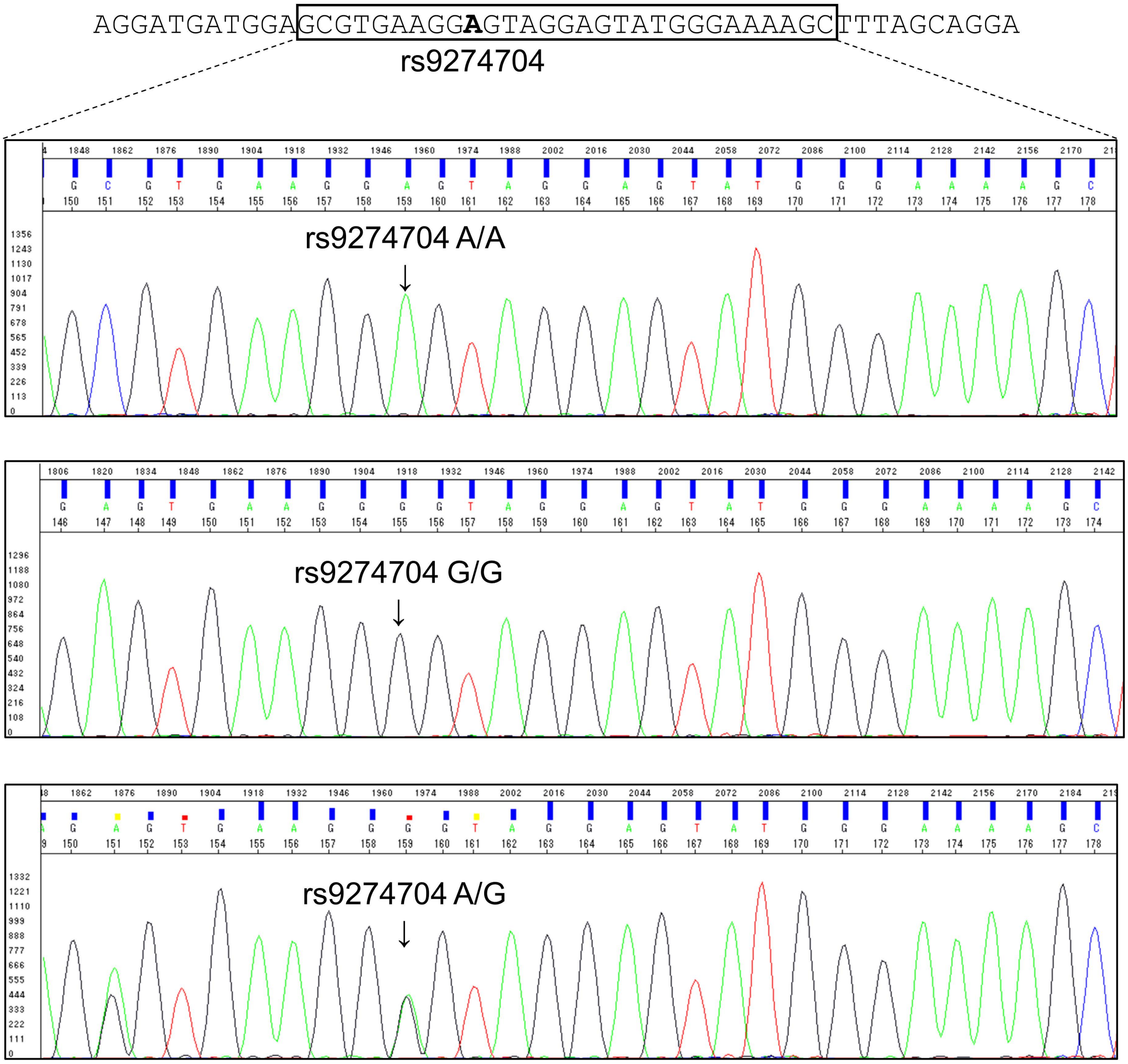

AGGATGATGGAGCGTGAAGGAGTAGGAGTATGGGAAAAGCTTTAGCAGGA
rs9274704
rs9274704 A/A
rs9274704 G/G
rs9274704 A/G

B

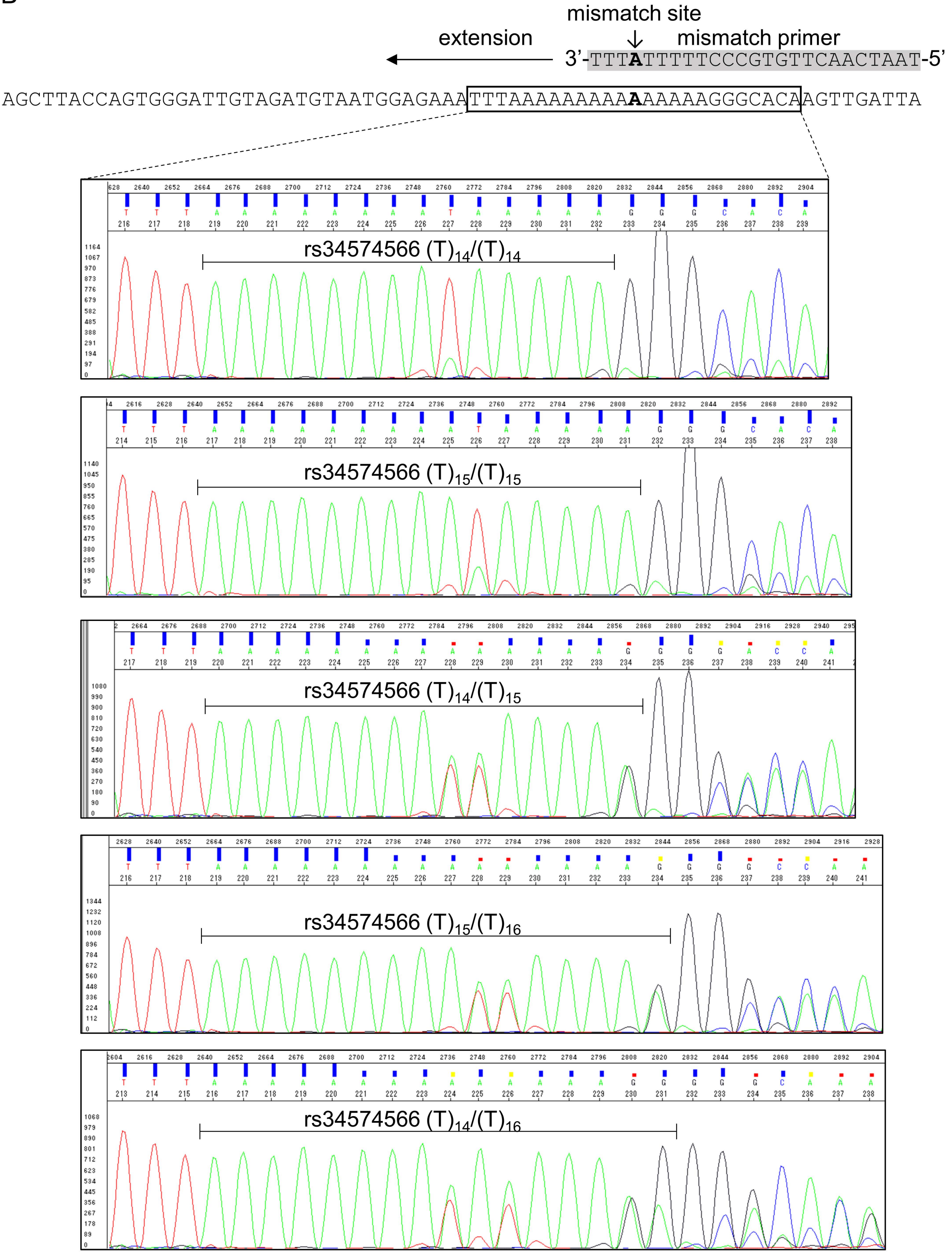
mismatch site
mismatch primer
extension
3'-TTTATTTTTCCCGTGTTCAACTAAT-5'
AGCTTACCAGTGGGATTGTAGATGTAATGGAGAAATTTAAAAAAAAAAAAAAAGGGCACAAGTTGATTA
rs34574566 (T)14/(T)14
rs34574566 (T)15/(T)15
rs34574566 (T)14/(T)15
rs34574566 (T)15/(T)16
rs34574566 (T)14/(T)16

**Supplementary Figure S1 Sequencing chromatograms of rs9274704 and rs34574566**

A. The nucleotide sequence of chr6:32,669,416-32,669,465(hg38, forward strand) was retrieved from the UCSC Genome Browser (https://genome.ucsc.edu/index.html). Representative chromatograms in individuals with rs9274704 A/A, G/G and A/G genotypes are shown.

B. The nucleotide sequence of chr10:9,000,766-9,000,834 (hg38, reverse strand) was retrieved from the UCSC Genome Browser. The primer is shaded in gray. A mismatched base (T→A) was introduced into the primer and is shown in bold. Representative chromatograms in individuals with rs34574566 $(T)_{14}/(T)_{14}$, $(T)_{15}/(T)_{15}$, $(T)_{14}/(T)_{15}$, $(T)_{15}/(T)_{16}$ and $(T)_{14}/(T)_{16}$ genotypes are shown.
